# Does diet contribute to socioeconomic differences in the prevalence of obesity among Indian adolescents?

**DOI:** 10.1101/2025.02.17.25322426

**Authors:** Neelam Kalita

**Author notes:** Corresponding author: Neelam Kalita.

## Abstract

**Background:** In India, the risk of obesity in adolescents between 10 and 19 years of age increases with socioeconomic advantage. Diet is a contributing factor to obesity, and obesity is a key risk factor for cardiovascular diseases (CVDs). However, the contribution of diet to socio-economic inequalities in overweight or obesity remains unclear.

**Aim:** To assess the mediating role of diet in the association between socioeconomic status (SES) and obesity among Indian adolescents.

**Data & Methods:** Data were obtained from the Comprehensive National Nutrition Dataset survey, a cross-sectional sample of the Indian adolescent population (N=35,830). Maternal educational attainment and household wealth index are used as proxies for SES. Diet quality was estimated using the Dietary Diversity Score (DDS) based on reported weekly intake of different categories of food. BMI for age Z-scores was used as a marker for obesity. Cross-section mediation was applied to estimate the proportions of obesity mediated by diet quality. Robustness checks were conducted by using alternative measures of diet quality.

**Results:** The log-odds of obesity were estimated to rise by 0.603 (95% CI: 0.334,0.926) and 0.714 (95% CI: 0.573, 0.865) in rural and urban adolescents, respectively, with higher levels of maternal education (post-secondary or above), compared to those from lower maternal education (secondary or below). In wealthier adolescents (from middle-, rich-, richest quintiles), the log-odds of obesity were estimated to rise by 0.710 (95% CI: 0.529, 0.891) and 1.325 (95% CI: 0.924, 0.1.726) in rural and urban adolescents, respectively, compared to poorer adolescents (from poorest and poor quintiles). The proportion of the association mediated by diet varied between <1% and over 4%.

**Conclusions:** The findings suggested that diet quality mediates socio-economic inequalities in obesity among Indian adolescents, but only a small proportion. Therefore, differences in diet quality might not be the primary route by which SES translates into differences in risk for obesity among Indian adolescents. Focussing on improving diet quality and other risk factors (behavioural, environmental, demographic, socio-cultural) could prevent obesity among Indian adolescents and, therefore, the future burden of CVD in India.

## 1 Introduction

Diet imbalance is a risk factor for overweight or obesity (henceforth referred as ‘obesity’), which is an intermediate risk factor for the development of cardiovascular diseases (CVDs).^1^ CVD is a leading cause of disease burden and mortality in India, accounting for over 80% of the disease burden and 28.1% of the total deaths among Indians, respectively.^2–4^ The social determinants of health, represented by socioeconomic status (SES), contribute to the CVD burden via a constellation of biological, behavioural and psychosocial risk factors.^5^ And, diet quality varies by SES among Indian adolescents, aged between 10 and 19 years.^6^

Adolescence is a critical period for the development of later health and disease as it bridges the nutrient deficits suffered during childhood and lays the foundation for healthy dietary habits for later life.^7^ ^8^ With 253 million, India has the largest adolescent population worldwide.^9^ Rapid socio-economic transition and urbanisation have led to a shift in dietary habits towards unhealthy choices and patterns, such as prepared and/or processed foods high in fat, sugar, and salt (HFSS). Among Indian adolescents, there are positive SES gradients in the prevalence of obesity and the consumption of food comprising fats and oils, sugar and jaggery and HFSS including junk food, fried food, sweets, and aerated drinks. More socio-economically advanced adolescents, measured by higher levels of maternal education attainment and household wealth quintiles, have higher consumption of these *‘unhealthy’* foods. This shift in dietary pattern may explain the socio-economic patterning of obesity, which is mostly concentrated among those from high SES and urban areas.^6^ To prevent the growing trend of obesity, there is a public health emphasis on improving the overall diet quality in early life-phases as it plays an important role in the development of many future health problems, including CVDs.^3^ ^10^

Previous findings, from studies based in India and globally, suggest a social gradient in the prevalence of obesity and associated CVD biomarkers including hypertension, diabetes, lipid anomalies: socially disadvantaged people are at higher risk of obesity and CVD biomarkers compared to those more privileged.^2^ ^11^ Those examining the role of diet in the association between SES and obesity reported wide variations in the proportion mediated, ranging from 0% to nearly 40%.^2^ ^12^ ^13^ However, these studies were conducted in high-income settings and focussed on adult populations. Therefore, the findings may not be generalisable to the current context given the significant differences in economic, social, physiological, and cultural factors that impact dietary behaviour. To date, few studies have examined whether the association between SES and obesity is moderated by diet in the Indian context and among adolescents. Therefore, the current study aimed to address this gap by examining whether diet mediated the association between SES and obesity, among Indian adolescents.

## 2 Materials and Methods

Data from India’s nationally representative Comprehensive National Nutrition Survey (CNNS), conducted in 2016-2018 among children aged 0 to 9 years and adolescents aged 10 to 19 years in the Indian population was used. The CNNS collected data on contemporaneous measurement of current diet (e, g., individual dietary intake using daily and seven-day food frequency questions of key food items), outcome of interest (e.g., health status and anthropometry), alongside information on socio-economic and demographic factors (e.g., maternal education, household wealth index, environmental conditions, etc.) from a large sample of adolescents (N=35,830). For adolescents aged 10-14 years, a parent was asked to be present during the interview to help the adolescent respond. For all the adolescents (10-19 years), parents answered questions about parental- and household factors (e.g., education level, sanitation).^14^ Description of the study variables is presented in Table 1.

**Table 1.** Description of study variables.

| Variable | Description | Statistical analyses |
| --- | --- | --- |
| **Outcome variable** |  |  |
| Overweight or obesity | BMI for age Z-scores (continuous, calculated using measured height and weight) | Binary: 0=Normal or underweight (Z-scores $\leq 1$) ; 1: Overweight or obese (z-scores $>1$)<br>Continuous (sensitivity analysis) |
| **Mediator variable** |  |  |
| DDS | Estimated from the frequency of consumption of food groups (obtained from FFQ collected in the CNNS) | Continuous; DDS scores ranging from 0 to 17; a score of 17 representing full compliance to dietary guidelines whereas 0 representing no compliance |
| **Exposure variable** |  |  |
| Mother's education level | 0 to 24 years of education, Continuous | Binary: 0= Completed secondary education or less (0-12 years); 1= post-secondary education or above<br>Continuous, same variable used (sensitivity analyses) |
| Wealth index | Categorical: 1=Poorest, 2=Poor, 3=Middle, 4=Rich, 5=Richest | Binary: 0=quintiles 1 & 2. 1= quintiles 3,4 & 5<br>Categorical, same variable used (sensitivity analyses) |
| **Covariates** |  |  |
| Sex | Binary, 1=Boy, 2=Female | Binary, 0=Female; 1=Male |
| Age | 10 to 19 years, Continuous | Continuous |
| Adolescent literacy based on school attendance | Binary: 1=Yes, 2=No | Binary: 0=No; 1=Yes |
| Religion | Categorical: 1=Hindu, 2=Muslim, 3=Christian, 4=Sikh, 5=Buddhist, 6=Other | Same variable used |
| Region | Categorical: 1=North, 2=Central, 3=East, 4=North-East, 5=West, 6=South | Same variable used |
| Caste | Categorical: 1=Scheduled caste, 2=Scheduled tribe, 3=Other Backward Caste, 4=General, 5=Don't know | Same variable used |

### 2.1 Socioeconomic Status

SES was defined by maternal education attainment and wealth index. Separate analyses are conducted for each of these variables. Following the approach adopted by a Canadian study by Nejatinamini et al.^15^ maternal education attainment was dichotomised into i) up to secondary (12 years or less of education), and ii) post-secondary (>12 years of education); wealth index into i) lower (quintiles 1 and 2), and ii) higher (quintiles 3, 4 and 5).

### 2.2 Diet Quality

Diet quality was estimated using Dietary Diversity Score (DDS), based on the number of food groups consumed per week. DDS assesses the adequacy and quality of diet using a simplified method.^16^ This measure was used for two reasons: i) lack of detailed dietary data including portion sizes, number of servings, and recipes required to estimate Diet Quality Index, and ii) previously used to assess diet quality in resource constrained settings and its association with CVDs;^16–22^ In the CNNS, dietary intake was measured using daily and seven-day food frequency questions on consumption of 17 food items, including cereals, dairy, pulses or beans, roots or tubers, dark green leafy vegetables, other vegetables, fruits, eggs, fish, chicken or meat, nuts or seeds, fats or oils, sugar or jaggery, fried food, junk food, sweets, and aerated drinks.^23^ Of the 17, 13 nutrient-rich food and recommended by ICMR^24^ and FAO,^25^ was given a score of 1 for each intake and counted only once even if it was consumed more than once per week. For the remaining 4 items high in fats, salt, and sugar contents (HFSS), the scoring was reversed: 1 for no intake and 0 for intake and counted only once even if it was consumed more than once per week. The reverse scoring pattern, based on the approach adopted by Yannakoulia et al.,^26^ was used to align with the guidance from the Indian Academy of Paediatrics Guidelines on the Fast and Junk Foods, Sugar Sweetened Beverages, Fruit Juices, and Energy Drinks.^27^ The scores ranged from 0 to 17; 17 representing full compliance to dietary guidelines and 0 no compliance.

Another measure, Food Variety Score (FVS), was also explored. This approach, adapted and modified from the methods described by Clausen et al.,^28^ Zainal Badari et al.,^29^ and Nithya et al.^30^, is a scoring system that uses the frequency of the food items consumed. While DDS focuses on dietary diversity and is linked to overall diet quality and nutritional adequacy, FVS measures the variety of individual food items consumed thereby providing insight into dietary variety but not necessarily nutritional quality. Scoring for FVS were categorised into two groups: foods recommended by ICMR and FAO^1^ and a subset of the HFSS food basket^2^ which are nutrient-poor but energy-rich. Scores were assigned for both the groups ranging from 7, everyday per week; 6, six times per week; and so on till 0, less than once a week. Overall scores were calculated by summing the frequency scores.

### 2.3 Obesity

Overweight or obesity (henceforth referred to as obesity) was measured by BMI for age z-scores, defined as a score of > 1SD.^31^

### 2.4 Covariates

The covariates used were age, sex, school attendance, religion, region and caste. These were chosen based on the individual- and household- information provided in the CNNS dataset, the theory of social determinants of health that outline non-medical factors that influence health outcomes,^32^ alongside India-specific factors that influence the associations between SES, diet and obesity^33–35^ and commonly used in previous literature.^36^ ^37^

### 2.5 Statistical Analysis

Cross-sectional mediation analyses were performed to examine the role of diet in the association between SES and obesity among Indian adolescents by area of residence (rural/urban). Drawing from the CNNS, area of residence appeared to have a differential impact in the associations among SES, diet and obesity; no differential impact was evident by gender (Supplementary appendix: **Error! Reference source not found.**). The mediation model is expressed with the following set of equations:

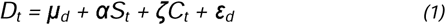

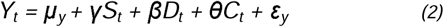

Y_t_ is the outcome- obesity, predicted by an independent variable - the exposure (maternal education and household wealth index), *S_t_*, and a mediator variable (diet), *D_t_*. α and β refer to unstandardised regression coefficients relating to *S_t_* and *D_t_*, and *D_t_* and *Y_t_* respectively. γ refers to the unstandardised regression coefficient relating *S_t_* and *Y_t_*, respectively when controlling for the mediator. μ*_d_* and μ*_y_* refer to the intercepts for the mediator and dependent variables, respectively, and ε refers to the random error associated with either the mediator or outcome effect (as specified in the subscript). *C_t_* is the list of the covariates (age, sex, adolescents attending school, religion, region, and caste). ζ and θ are the unstandardised regression coefficients relating to *C_t_* and *D_t_* and *C_t_* and *Y_t,_* respectively.^38^ The primary effects of interest in examining the mediation process are the direct and indirect effects, and the proportion of the total mediated effect. With reference to equations 1 and 2 above, γ refers to the direct effect. The indirect effect, which denotes how S_t_ influence Y_t_ through the mediating variable D_t_ is estimated as the product of α and β. Finally, the proportion of the total effect mediated by D_t_ is estimated by the following equation:

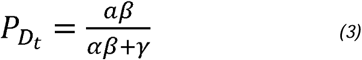

To establish the occurrence of mediation, the following conditions, outlined by Baron and Kenny^39^ and Judd and Kenny,^40^ are required: i) exposure is causally related to outcome, before mediator is added to the model; ii) exposure is causally related to mediator (i.e., α is non-zero); and iii) mediator is causally related to outcome, when controlling for exposure (i.e., β is non-zero). Causation was assumed based on these conditions. However, the data did not allow for any endogeneity or reverse causality, and the analyses only controlled for the observables.

The association between SES and DDS was assessed using linear regression, and that between DDS and obesity and SES and obesity using logistic regressions, adjusting for the covariates. Statistical model fits were analyses using residuals and Goodness-of-Fit statistics. To ascertain no unmeasured confounding for exposure-outcome, mediator-outcome, and exposure-mediator associations, the potential confounders were included as covariates for all the associations between the exposure, mediator, and outcome that were available from the CNNS dataset.

To assess robustness of the results, sensitivity analyses are conducted using i) different dietary measures as mediators including FVS, DDS score using 13 recommended food groups, and DDS using 4 HFSS food groups, and ii) maternal education and wealth as continuous variables. All analyses were conducted using Stata version 17 (Stata Corp, Texas, USA). P values < 0.05 were considered statistically significant.

### 2.6 Ethics

International and national ethical approvals were obtained from the Population Council’s Institutional Review Board in New York and Post Graduate Institute of Medical Education and Research in Chandigarh, respectively, prior to initiating the CNNS survey. All the respondents underwent an informed consent and assent process for participating in the survey: for 10-year-olds, informed consent was obtained from parent/caregiver; for 11-to-17-year-olds, from both parent/caregivers and adolescents; and for 18-to-19-year-olds, from adolescents. Further details on the sampling and methodology are described in detail elsewhere.^14^

## 3 Results

### 3.1 Characteristics of the Sample

Table 2 shows the characteristics of the included sample. In total, 32,120 participants were included in the analysis, of which 49% were girls and 51% were boys. The prevalence of obesity was highest among richest wealth quintile (15%) and higher level of maternal education (24%). The mean DDS scores higher among urban, wealthier quintiles and higher maternal education.

**Table 2.**
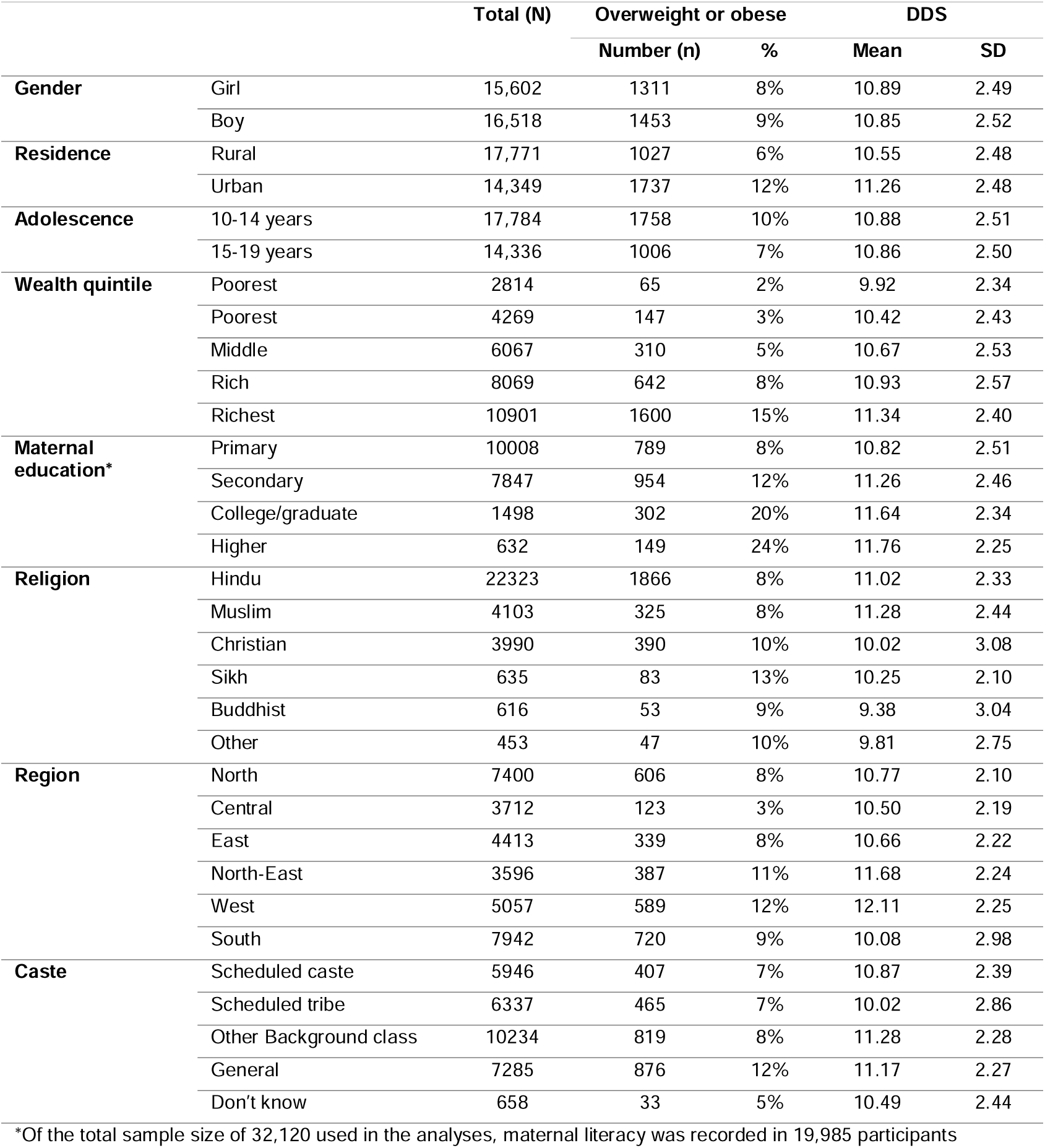
Description of the included sample, CNNS 2014-16.

### 3.2 Associations between SES, dietary measure, and obesity

Table 3 displays the covariates adjusted associations between exposure-mediator, mediator-outcome, and exposure-outcome, by area of residence. Full regression results are in supplementary appendix. Overall, the results indicate positive trends in these associations for both rural and urban adolescents. Briefly illustrating the results using the estimates from rural adolescents, those with higher maternal education (post-secondary and above) are likely to have high DDS by 0.279 points, on average, compared to those with lower maternal education (secondary and below); the DDS score also increases by 0.362 for those from a wealthier household (middle, rich, richest quintiles) compared to those from poorer households (poorest, poor quintiles). Similarly, the log-odds of obesity is estimated to rise by 0.016 with one point increase in DDS. There is also a positive association between SES and obesity.

**Table 3.** Associations between exposure-mediator, mediator-outcome, exposure-outcome.

|  | *Coefficient (SE)* | *95% Confidence Interval* | *p-value* |
| --- | --- | --- | --- |
| **Rural** |  |  |  |
| Maternal education | 0.279 (0.117) | (0.049, 0.509) | 0.017* |
| Wealth index | 0.362 (0.041) | (0.282, 0.441) | 0.000* |
| **Urban** |  |  |  |
| Maternal education | 0.358 (0.061) | (0.239, 0.478) | 0.000* |
| Wealth index | 0.439 (0.084) | (0.274, 0.605) | 0.000* |
| **Association between mediator (DDS) and outcome (Obesity)** |  |  |  |
|  | *log-odds (SE)* | *95% Confidence Interval* | *p-value* |
| **Rural** |  |  |  |
| DDS | 0.016 (0.014) | (-0.011, 0.044) | 0.238 |
| **Urban** |  |  |  |
| DDS | 0.020 (0.012) | (-0.003, 0.042) | 0.090 |
| **Association between exposure (SES) and outcome (Obesity)** |  |  |  |
|  | *log-odds (SE)* | *95% Confidence Interval* | *p-value* |
| **Rural** |  |  |  |
| Maternal education | 0.630 (0.151) | (0.334, 0.926) | 0.000* |
| Wealth index | 0.710 (0.092) | (0.530, 0.891) | 0.000* |
| **Urban** |  |  |  |
| Maternal education | 0.714 (0.072) | (0.573, 0.854) | 0.000* |
| Wealth index | 1.325 (0.205) | (0.924, 1.726) | 0.000* |

Data source: CNNS 2016-2018
\*Statistically significant at $p < 0.05$

### 3.3 Mediation of diet in the SES-obesity association

Table 4 shows the results of the cross-section mediation analysis by area of residence and discussed below. Overall, these indicated that improved SES leads to better diet quality (measured by DDS); but diet quality had a minimal effect on obesity among Indian adolescents.

**Table 4:** Results from cross-section mediation analysis.

|  | Coefficients | 95% Confidence Intervals |  | p-value |
| --- | --- | --- | --- | --- |
|  |  | Lower | Higher |  |
| RURAL |  |  |  |  |
| Direct effects of SES on obesity |  |  |  |  |
| Maternal education ->DDS->obesity | 0.625 | 0.329 | 0.921 | 0.000* |
| Wealth ->DDS->obesity | 0.707 | 0.526 | 0.887 | 0.000* |
| Indirect effects of SES on obesity through DDS |  |  |  |  |
| Maternal education ->DDS-> obesity | 0.004 | -0.007 | 0.015 | 0.504 |
| Wealth ->DDS->obesity | 0.004 | -0.005 | 0.013 | 0.412 |
| Total effects of SES on obesity |  |  |  |  |
| Maternal education ->DDS ->obesity | 0.630 | 0.334 | 0.926 | 0.000* |
| Wealth -> DDS ->obesity | 0.710 | 0.529 | 0.891 | 0.000* |
| **URBAN** |  |  |  |  |
| **Direct effects of SES on obesity** |  |  |  |  |
| Maternal education ->DDS ->obesity | 0.708 | 0.568 | 0.849 | 0.000* |
| Wealth ->DDS ->obesity | 1.318 | 0.917 | 1.719 | 0.000* |
| **Indirect effects of SES on obesity through DDS** |  |  |  |  |
| Maternal education ->DDS -> obesity | 0.005 | -0.005 | 0.015 | 0.312 |
| Wealth ->DDS ->obesity | 0.007 | -0.004 | 0.018 | 0.191 |
| **Total effects of SES on obesity** |  |  |  |  |
| Maternal education ->DDS ->obesity | 0.714 | 0.573 | 0.854 | 0.000* |
| Wealth -> DDS ->obesity | 1.325 | 0.924 | 1.726 | 0.000* |

\*Statistically significant as $p < 0.05$ level

#### Impact of maternal education on obesity via diet

- ***Rural adolescents:*** The log-odds of obesity was estimated to rise by 0.630 (95% Confidence Interval(CI): 0.334, 0.926) with increase in maternal education. Of this total effect, the direct effect that was not mediated by DDS was 0.625 (95% CI: 0.329, 0.921). The indirect effect, which was mediated by DDS, was 0.004 (95% CI: -0.007, 0.015) although this was not statistically significant. Therefore, the proportion of the total effect of maternal education on obesity that was mediated by DDS was 0.63%.
- ***Urban adolescents:*** The total effect of maternal education on obesity was 0.714 (95% CI: 0.573, 0.865), meaning the log-odds of obesity increased with increase in maternal education. The direct effect that was not mediated by DDS was 0.708 (95% CI: 0.568, 0.849) and the indirect effect, mediated by DDS, was 0.005 (95%CI: -0.005, 0.015). This implies that the proportion mediated by DDS was marginal at 0.70%.

#### Impact of household wealth on obesity via diet

- ***Rural adolescents:*** Those from wealthier households were likely to have higher risk of obesity with the log-odds estimated to rise by 0.710 (95% CI:0.529, 0.891), compared to those from poorer households. Of this total effect, the direct effect was 0.707 (95% CI: 0.526, 0.887). The indirect effect, mediated by DDS, was 0.004 (95% CI:-0.005, 0.013), indicating that the proportion mediated by DDS was 0.56%.
- ***Urban adolescents:*** Like their rural counterparts, urban adolescents from wealthier households were more likely to be obese with the log-odds estimated to rise by 1.325 (95% CI: 0.924, 1.726) compared to those from poorer households. Most of this total effect was not mediated via DDS; the direct effect was 1.318 (95% CI: 0.917, 1.719); whereas the indirect effect was 0.007 (95% CI: -0.004, 0.018) indicating a minimal proportion of 0.53% that was mediated via DDS.

### 3.4 Robustness checks

Results of the robustness checks are presented in Supplementary Tables 3-4. Compared to DDS, applying FVS_13 food groups_ as mediator explained a higher proportion of the associations between SES and obesity for both rural and urban adolescents. It explained 2.54% (for rural) and 4.90% (for urban) of the associations between maternal education and obesity; and 2.54% (for rural) and 4.15% (for urban) of the associations between wealth index and obesity, respectively. Applying FVS_4 HFSS food groups_ explained 2.06% and 1.12% of the associations between maternal education and obesity in rural and urban adolescents; 2.68% and 1.96% of the associations between wealth index and obesity in the two populations, respectively. Decomposing the overall DDS into two scores (DDS_13 food groups_ and DDS_4 HFSS food groups_) shows higher proportion of association being explained by the two decomposed scores for the association of SES and obesity, compared to the overall DDS used in the base case. The proportions explained by these scores varied between 1.43% and 3.10% for rural adolescents and between 1.96% and 2.72% for urban adolescents. Using the SES as continuous variables showed a very minimal mediating effect of DDS in their association on obesity. Overall, the directions of the results were similar to the base case; however, the magnitude of the mediating effect varied depending on the diet quality measure used in the analyses.

## 4 Discussion

In this sample of adolescents in the Indian population, the risk of obesity was found to be positively associated with improvement in SES; however, diet quality did not fully explain this association. This suggests that whilst higher consumption of certain nutrient-poor, energy-dense foods including fats & oils, sugar and jaggery and HFSS among well-off adolescents may contribute to obesity, a host of other factors such as lack of physical activity, increased use of technology and screen-time, parental educational aspirations, etc. may play an important role.^6^ ^41^ ^42^ The current findings contradict those in the literature based on high-income countries where the evidence suggests those from lower SES (e.g., lower education levels) are more likely to have poorer diet quality.^2^ ^43^ ^44^

In the analyses, diet quality explained some of the association between SES and obesity, with the proportion varying between <1% and over 4%, depending on the diet quality measure (DDS versus FVS) used. Whilst both the dietary measures assess the variety of food consumed, DDS focuses on dietary diversity and linked to overall diet quality and nutritional adequacy, whereas FVS provides an insight into dietary variety and not necessarily nutritional quality.

Comparing these findings to the literature is limited by- different contexts (HIC versus LMIC); differing analytical mediation methods, -measures of overweight or obesity (self- reported or BMI derived), and most importantly, the way diet quality is assessed.^15^ ^43^ Furthermore, the existing literature highlighted wide variation in the proportion of association between SES and obesity mediated by diet. For example, a Swiss study found diet mediated between 22.1% and 35.6% of the association between education and obesity, depending on the obesity marker used.^2^ On the contrary, an Australian study reported no mediation of educational inequalities in BMI using a “healthy take-away foods” index whereas an “unhealthy take-away foods” index explained 15% of the mediation.^12^ Another European study found vegetables intake explained 8% of the association between neighbourhood SES and BMI, whilst fruit, soft drinks, and sweets intake didn’t explain any part of the association.^13^

### 4.1 Implications for Public Health

Continuous expansion of obesity together with its detrimental effects on CVDs and long-term disease burden, characterised by a higher relative risk, earlier age of onset, higher fatality and premature deaths, highlight an immediate need for improvements in diet quality among young population in India, alongside improvements in other behavioural factors.^45^ While our findings may suggest that equalising diet differential might not have much impact on the SES gradient in obesity, our results need to be treated with caution due to the inherent limitations on the nature of the data available. Despite the current findings, there is a need to strive for improvements in diet quality among Indian adolescents to tackle the double burden of undernutrition and overnutrition. This can be achieved by incorporating major structural changes to transform public awareness and current food environments into those that facilitate and promote healthy eating behaviours, which in turn, would pave the way for a population-wide shifts at the macro level towards healthier diets. Another approach would be to have targeted interventions aimed at the adolescents, which again are likely to benefit not only the young population but also the wider population in the long run.^2^ ^46–50^

Our study did not ascertain the association between SES and CVD biomarkers (including, HbA1c, Ratio of Total Cholesterol to HDL-c, total triglycerides, and hypertension) and therefore, the mediating role of diet. Nonetheless, diet remains a distal factor that influences these factors indirectly through its impact on obesity. Epidemiologically, it is hypothesised that adolescents consuming unhealthy diets are more likely to suffer from lipid anomalies which are a significant covariate of CVD. As illustration, 77% of Indian adolescents who consumed unhealthy diets suffered from lipid anomalies (non-HDL cholesterol and LDL cholesterol); the burden was highest among overweight or obese adolescents, followed by pre-diabetic and those with micronutrient deficiencies.^51^ Furthermore, there is an association between overweight or obesity and a higher prevalence of high blood pressure among Indian adolescents.^52^ This highlights the need to monitor high blood pressure from early adolescence because the trajectory curves from childhood to adulthood indicate higher blood pressure in childhood progresses to hypertension in young adulthood.^52–54^

### 4.2 Study strengths and limitations

The key strength of our study is that it is a pioneer in investigating the mediating effects of a modifiable risk factor for obesity in the Indian context. Secondly, our study used a nationally representative sample of Indian adolescents using the CNNS dataset, the first in the country to provide data on anthropometry (for 5-14 years), micronutrient deficiencies and risk factors for non-communicable diseases (15-19 years) for children and adolescents.^14^ To our knowledge, no previous studies have examined the mediating role of diet in influencing the impact of SES on obesity among Indian population. Third, a key strength of the current sample lies in the comprehensive dietary intake measurements using 24h dietary recalls, and the use of objectively measured markers of overweight and obesity. Fourth, the sample size was optimal as it reflected pan-India, including all the regions of the country.^2^ Finally, wide range of robustness checks were conducted to address any uncertainties.

Our study has several limitations. The main limitation is the cross-sectional nature of the data, which does not allow for temporal examination of exposure, mediator, and outcome and hence, causation. The magnitude of the relationships may be biased as we controlled only on observables, and did not allow for any endogeneity or reverse causality. Nonetheless, the consistent associations found in the analysis (between exposure-mediator, mediator-outcome, and exposure-outcome) agree with an extensive literature linking diet quality to obesity and socioeconomic conditions, thereby reiterating validity of the findings. Secondly, the biases associated with cross-section mediation analysis due to intermediate confounding and information bias are incorporated within the current analyses. Cross-section mediation is specific to linear models and doesn’t explore non-linear relationship between the exposure and mediator, unlike flexible method as counterfactual mediation using g-computation that can handle any combination of binary and continuous variables, incorporate exposure-mediator interactions and intermediate confounders. Thirdly, the mediation analyses were conducted based on the fundamental assumption of causality wherein the exposures were assumed to occur first. It is acknowledged that while this may hold for maternal education levels, there is uncertainty whether the assumption holds for household wealth index. Because the CNNS collects information on the current household items owned by the family, which informs the estimation of the household wealth index. The survey does not provide any information on the historic household income and wealth status. Therefore, the findings using household wealth index as exposure should be treated with caution. Fourth, dietary behaviour is a behavioural risk factor that often occurs with other behavioural choices such as levels of physical activity and smoking. Those adopting healthy dietary choices are usually more likely to be physically active and make healthy lifestyle choices (e.g. abstain smoking). It is, therefore, difficult to dissociate the specific effects of dietary components from levels of physical activity and other behavioural choices, as smoking, outside the setting of a controlled trial.^55^ Fifth, while the mediation analyses included confounding factors as covariates, there may be other factors such as lifestyle and other behavioural factors, parental occupations, family beliefs and behaviour, genetic factors affecting the association. These were not incorporated in the analyses as it is beyond the remit of the available dataset and available information for Indian adolescents. Sixth, our study could not estimate commonly used Diet Quality Index (such as, Youth Healthy Eating Index, Adolescent Micronutrient Quality Index, Diet Quality Index for Adolescents, Alternate Healthy Eating Index) to quantify the overall quality of an individual’s dietary intake due to lack of a detailed food composition database including portion sizes and recipes. However, in resource-constraint setting as India, DDS is more commonly used as the estimation of this straightforward measure does not require a food composition database.^56^ Finally, central adiposity- an important outcome for adolescents was excluded as an outcome; the inclusion of this measure could reveal further information.

## 5 Conclusions

The current findings suggest diet quality has a minimal mediating role in explaining the socio- economic inequalities in obesity among Indian adolescents. Differences in diet quality might not be the primary route by which SES translates into differences in risk for obesity among Indian adolescents. However, our findings are subject to several limitations. Focussing efforts on improving diet quality together with addressing other risk factors (behavioural, environmental, demographic, socio-cultural) could prevent obesity among Indian adolescents and therefore, the future burden of CVD in India.

## Data Availability

The Comprehensive National Nutrition Survey (CNNS) data are owned by the Ministry of Health and Family Welfare (MoHFW) of the Government of India. The MoHFW and the United Nations Children Fund, India Country Office, released the data used in this paper for public use. The analytic code for the analyses can be made available to the corresponding author upon request.

## Acknowledgements

The author thanks Professor Susan Griffin and Dr Sumit Mazumdar for commenting on earlier versions of the draft.

## Author contributions

The author conceived the idea, conducted all the statistical analyses, and drafted the manuscript. The author was involved at each iteration of drafting and approved the final paper. The author had access to raw data.

## Compliance with ethical standards

### Ethical approval

The CNNS was conducted according to the guidelines laid down in the Declaration of Helsinki, and all procedures involving human subjects were approved by the Population Council’s International Review Board (New York, USA) and ethics committee of the Post Graduate Institute of Medical Education and Research (Chandigarh, India). No separate ethical approval was required or taken for the secondary analyses conducted as part of this study.

## Footnotes

1 Includes 13 food groups: cereals; milk or milk products; pulses or beans; roots and tubers; green leafy vegetables; other vegetables; fruits; fats and oils; nuts and oilseed; sugar and jaggery; eggs; fish; chicken or meat

2 Includes 4 food groups: fried food, junk food, sweets and aerated drinks.

