## Supplementary appendix for "Does diet contribute to socioeconomic differences in the prevalence of obesity among Indian adolescents?"

### 1. Slope of DDS by gender and area

Figure 1 Associations of SES, DDS, overweight/obesity by gender and area of residence (based on CNNS 2016-18)

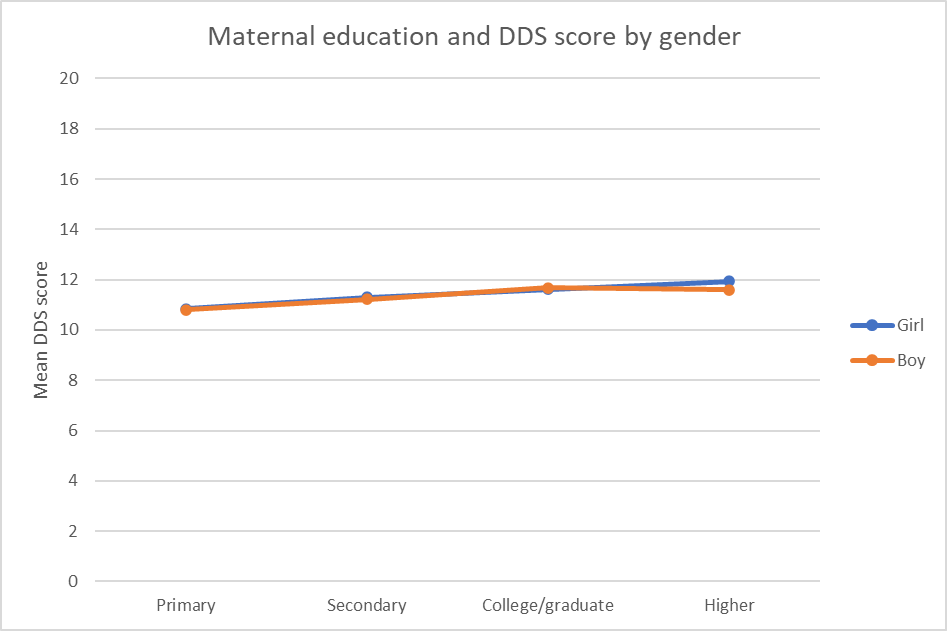

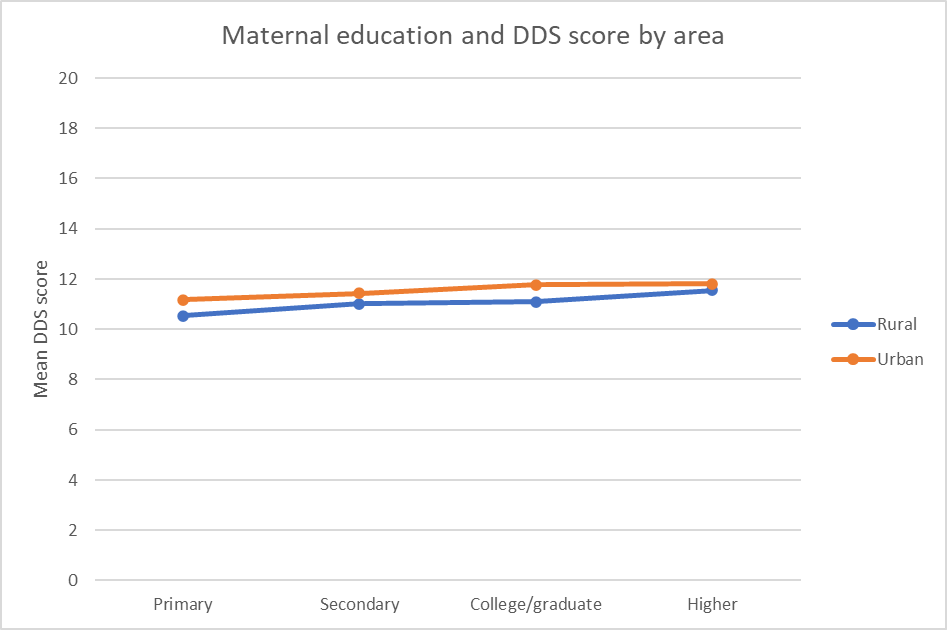

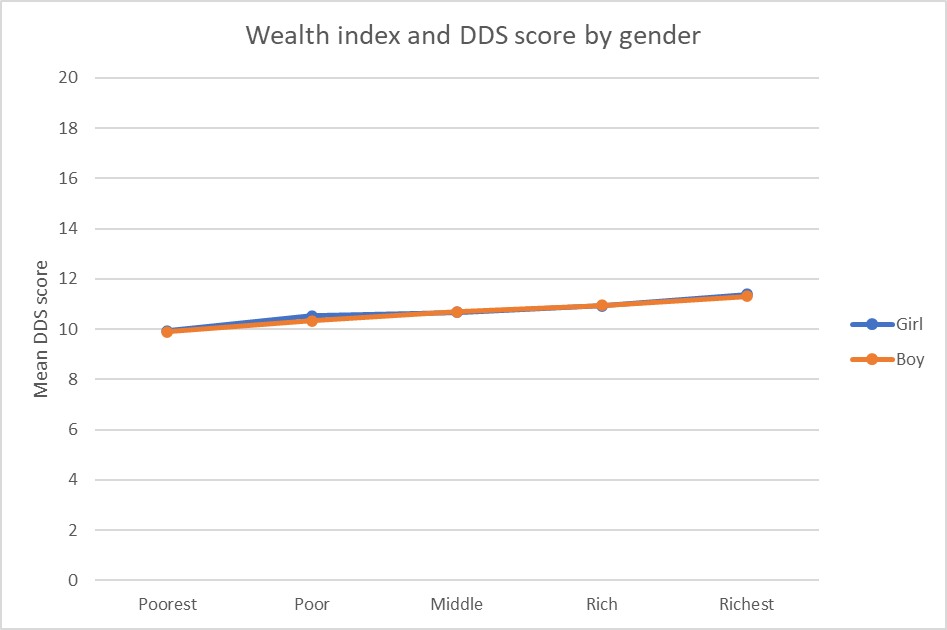

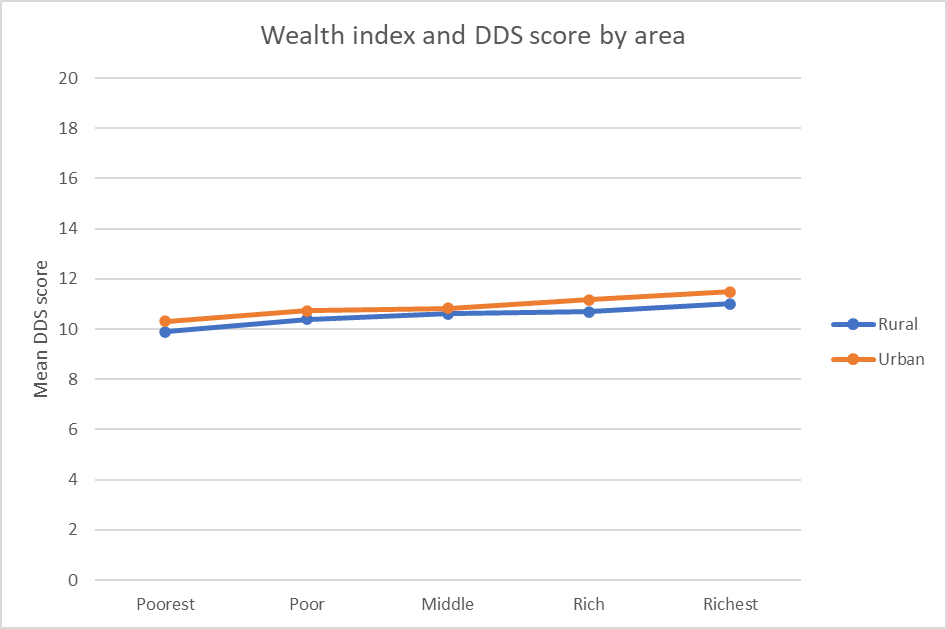

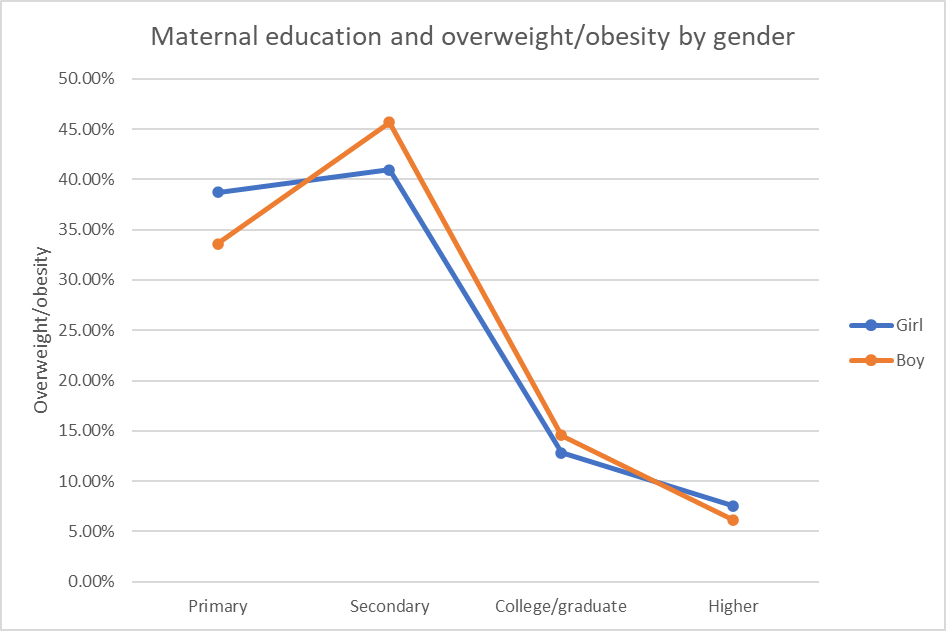

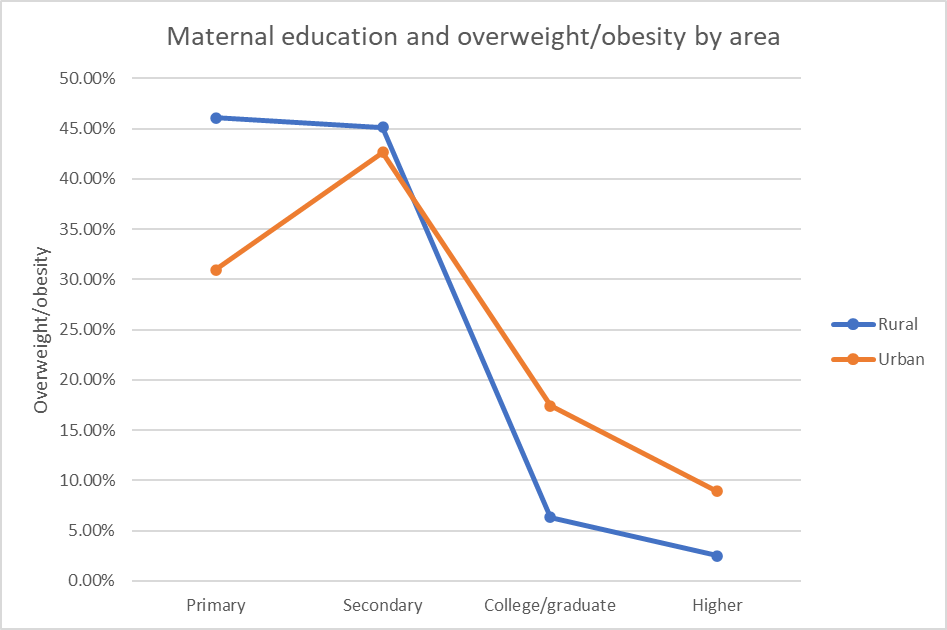

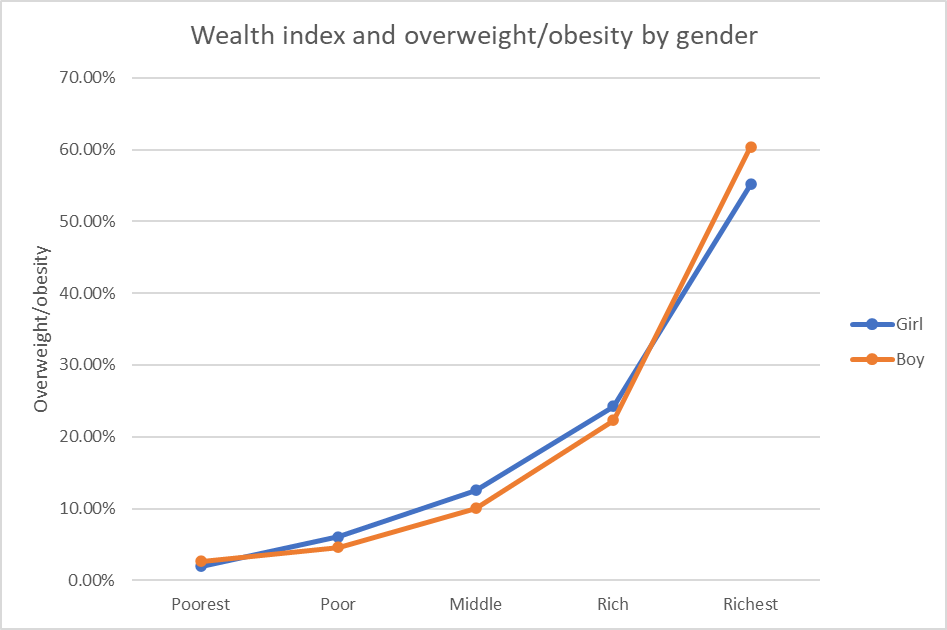

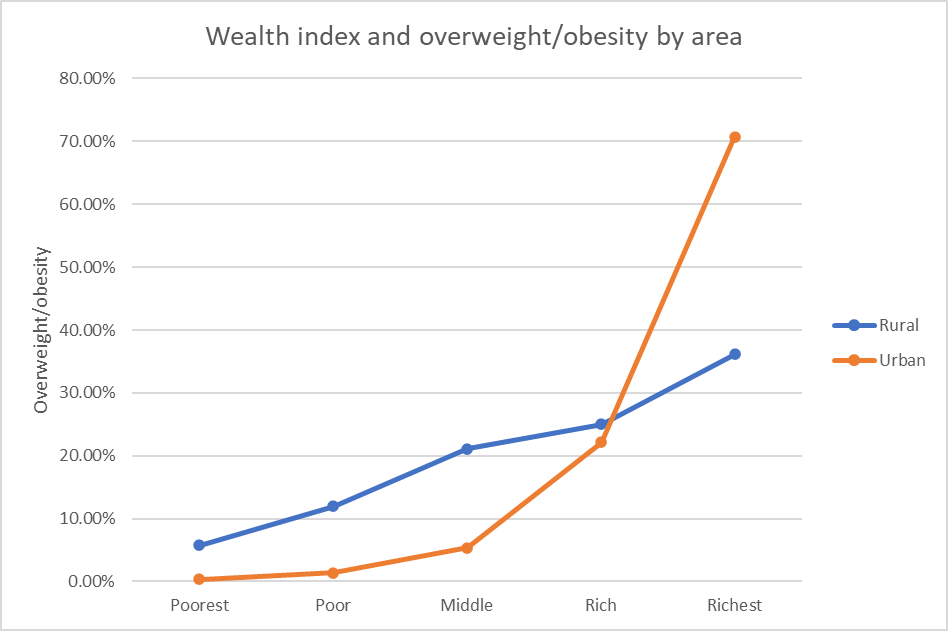

### 2. Association between SES, DDS and overweight/obesity

#### 2.1 **Rural**

##### 2.1.1 Association between maternal education and DDS

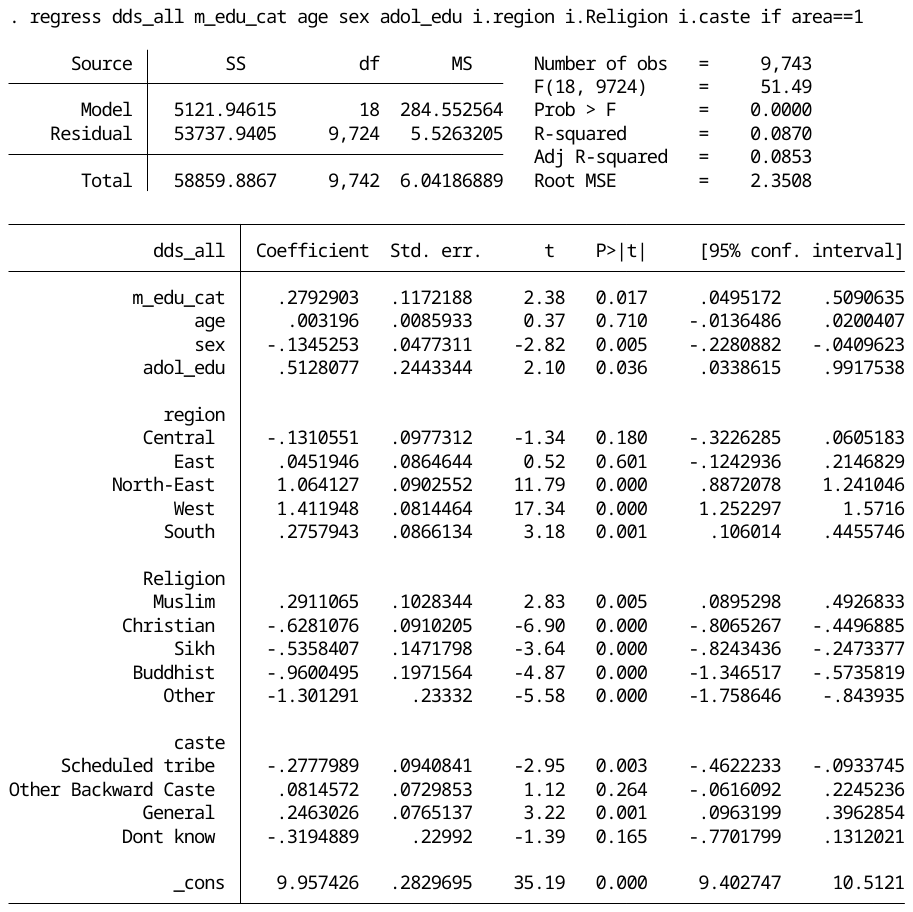

##### 2.1.2 Association between wealth index and DDS

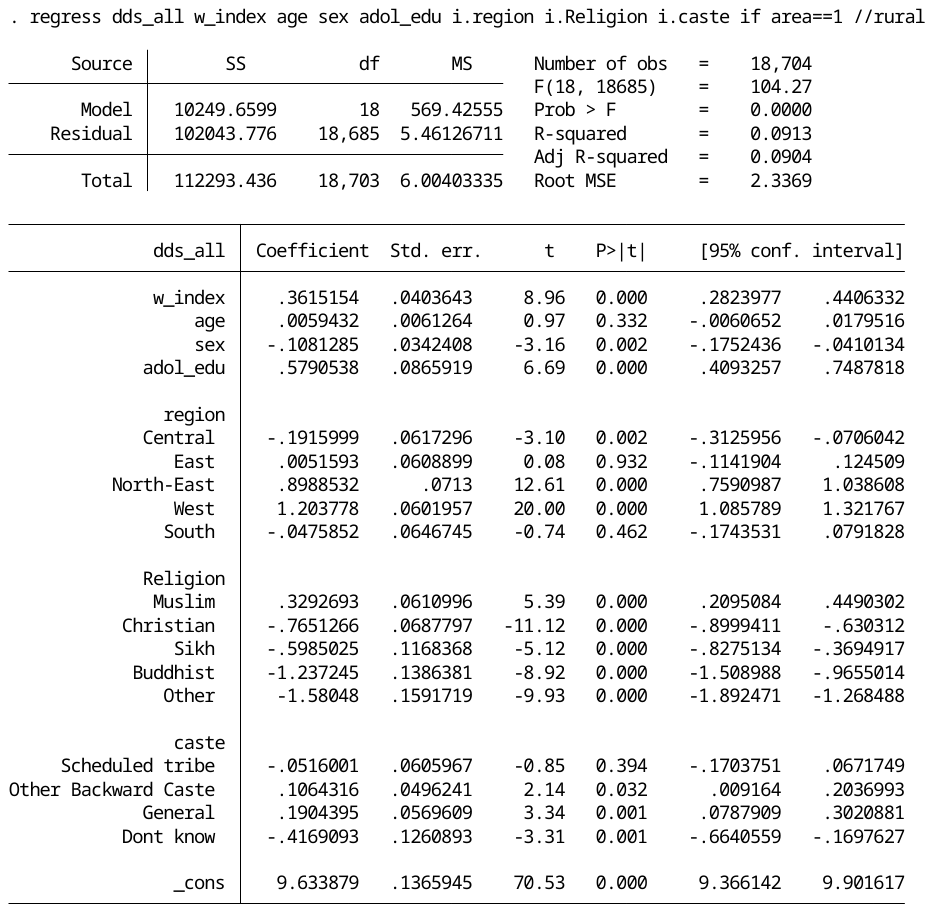

##### 2.1.3 Association between DDS and overweight/obesity

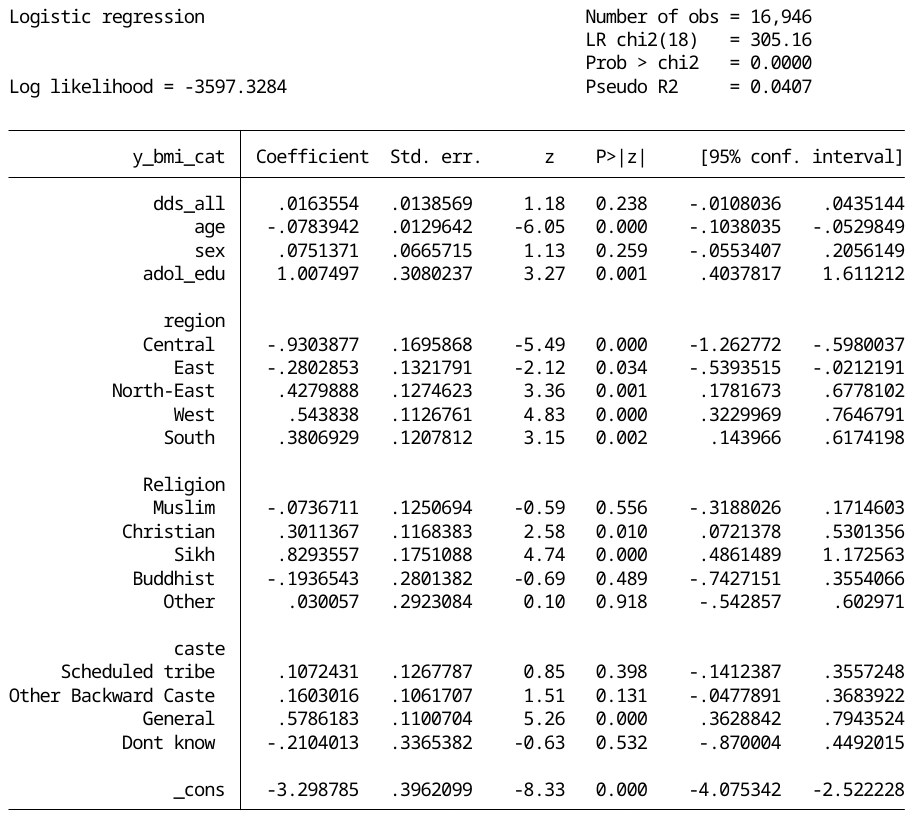

##### 2.1.4 Association between maternal education and overweight/obesity

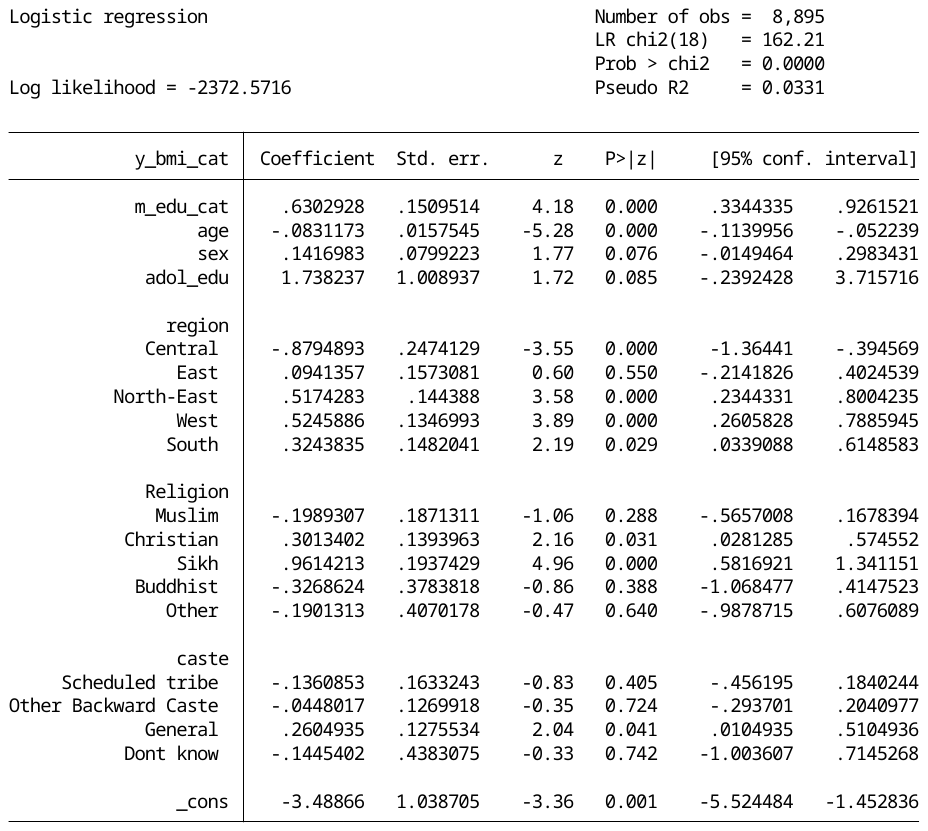

##### 2.1.5 Association between wealth index and overweight/obesity

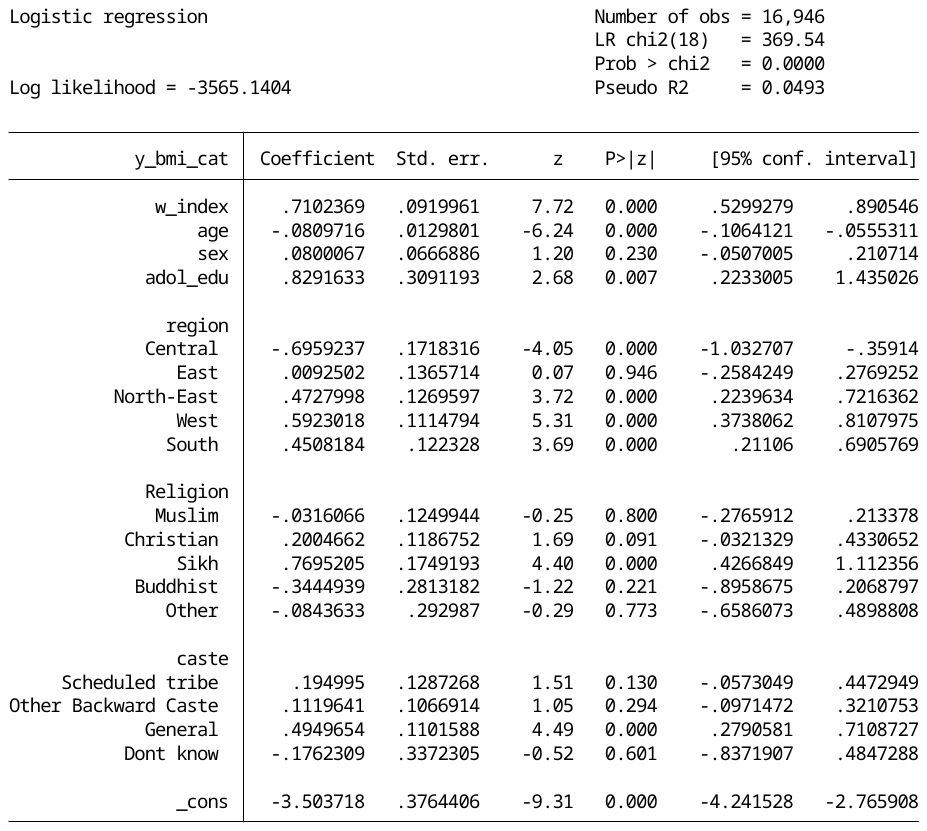

#### 2.2 **Urban**

##### 2.2.1 Association between maternal education and DDS

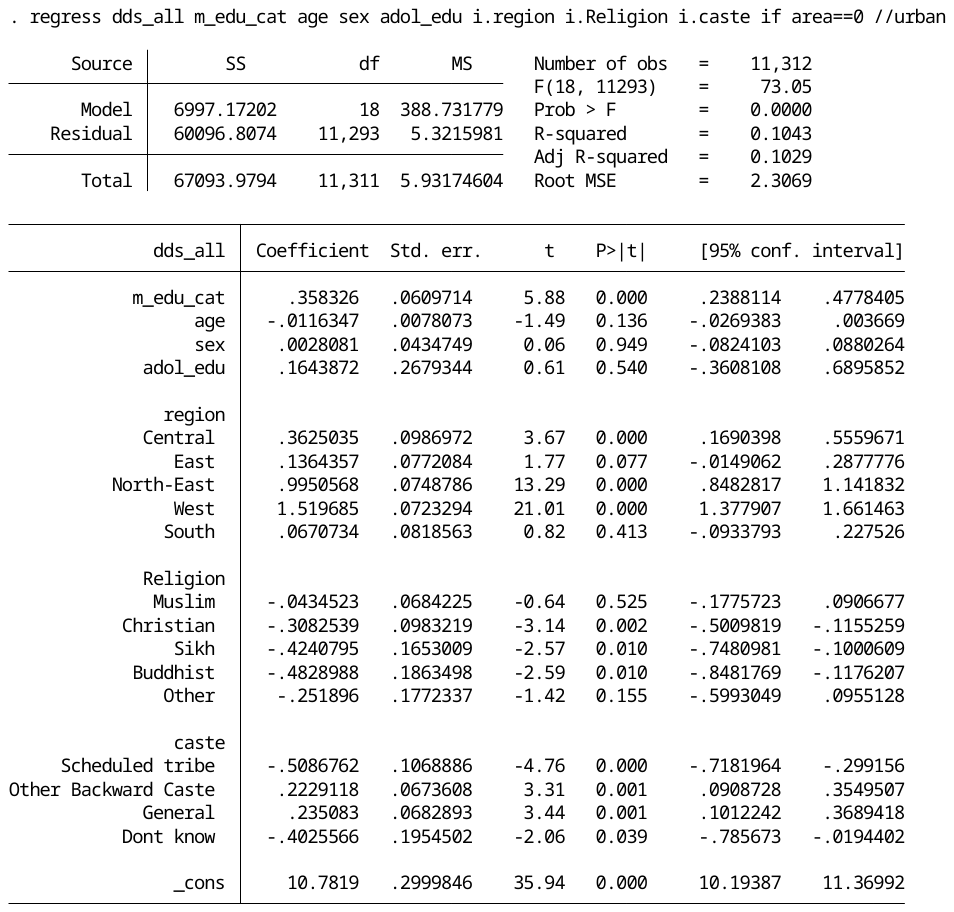

##### 2.2.2 Association between wealth index and DDS

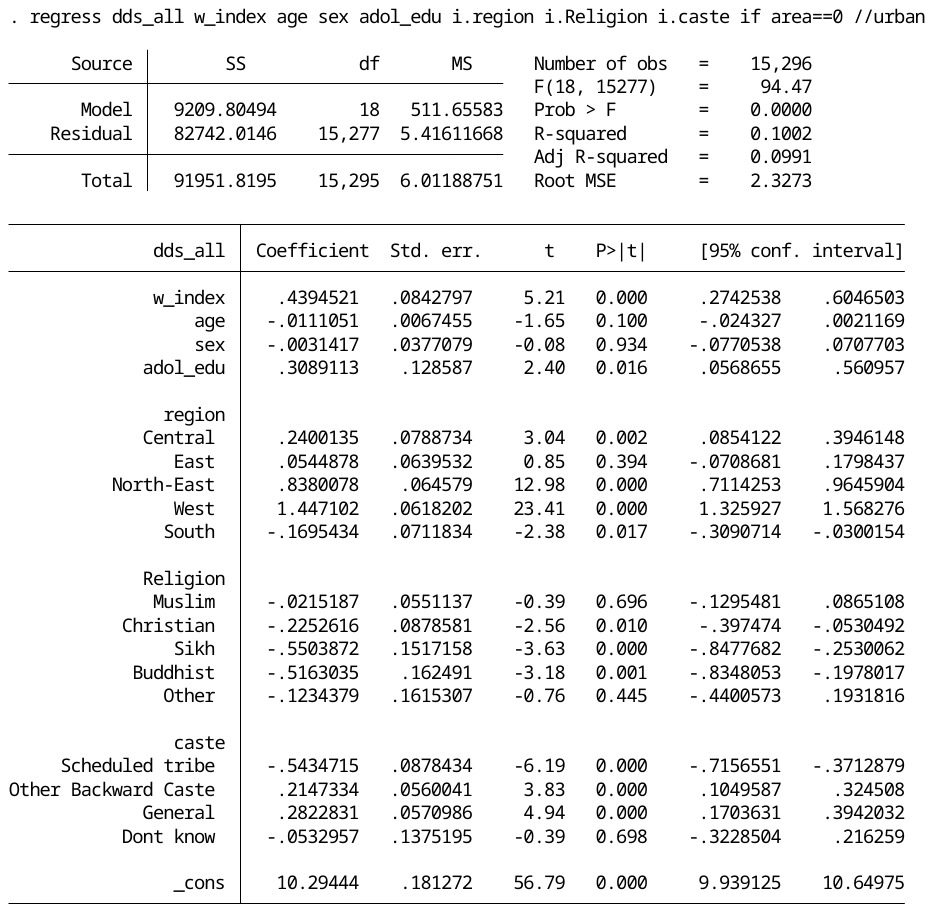

##### 2.2.3 Association between DDS and overweight/obesity

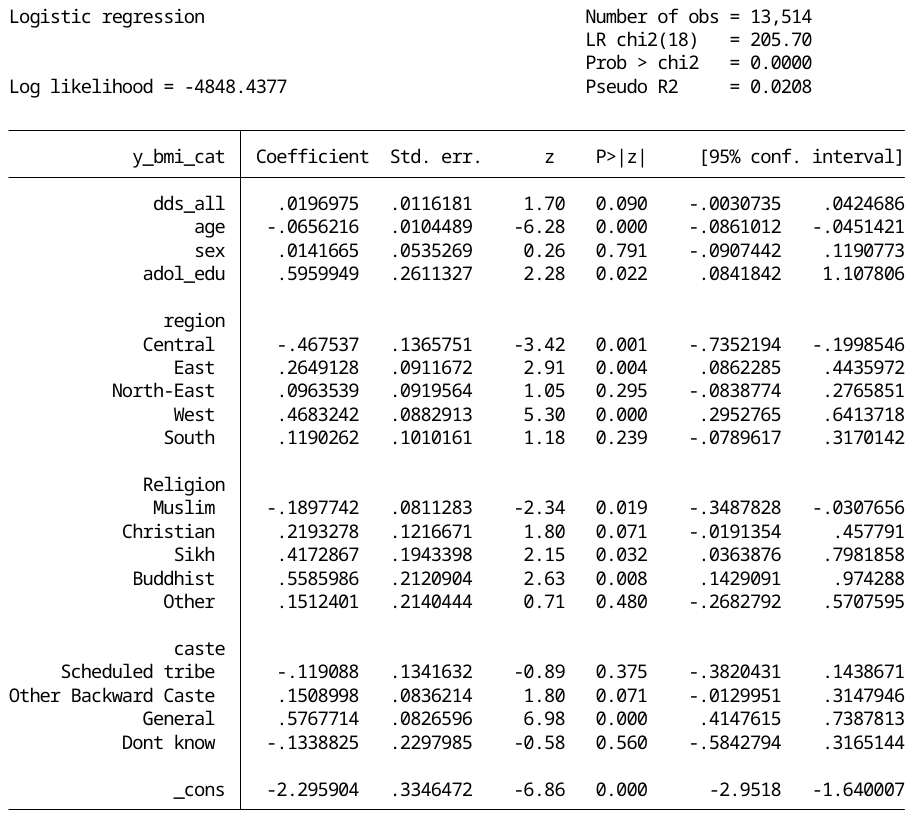

##### 2.2.4 Association between maternal education and overweight/obesity

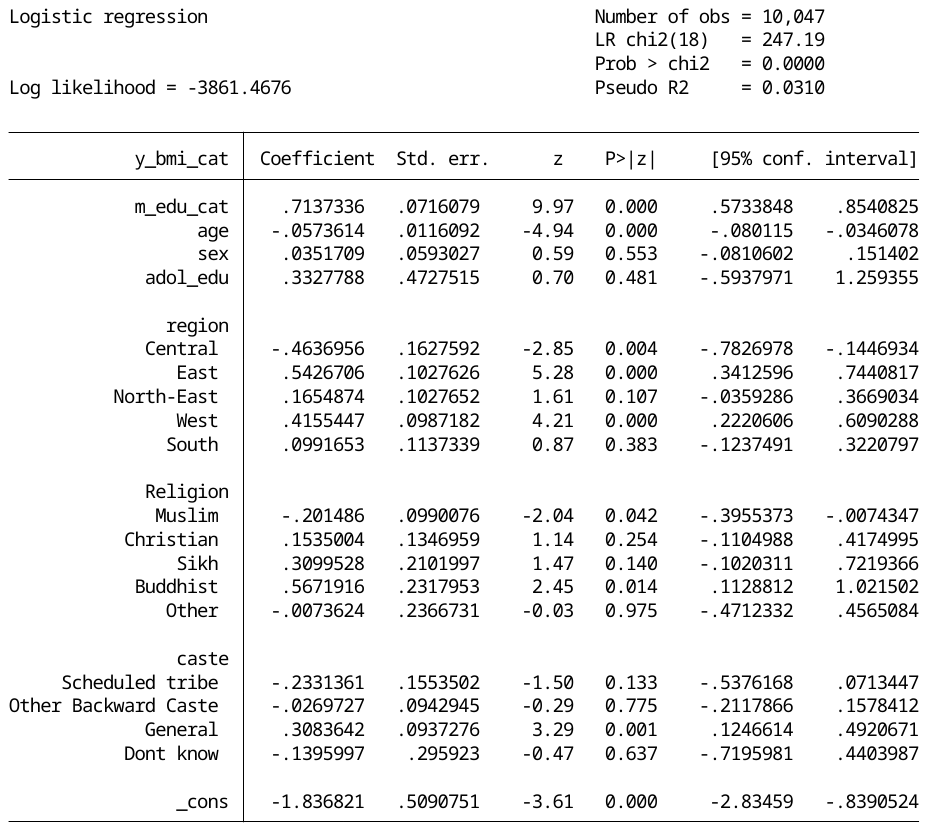

##### 2.2.5 Association between wealth index and overweight/obesity

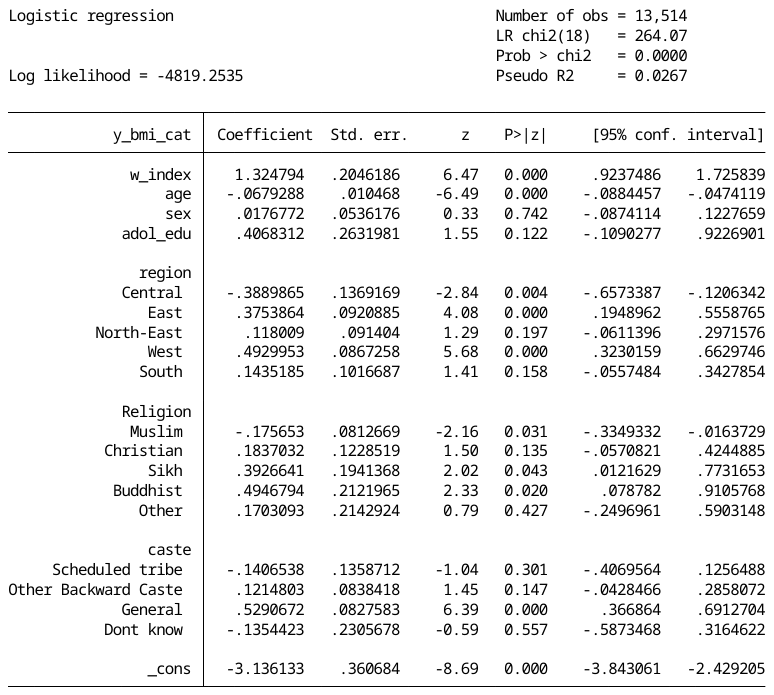

### 4. Cross section mediation

#### 4.1 **Rural**

##### 3.1.1 Maternal education – DDS- overweight/obesity

**Exposure: m_edu_cat (binary); Mediator: dds_all (continuous); Outcomes: y_bmi_cat (binary)**

**Total effect**

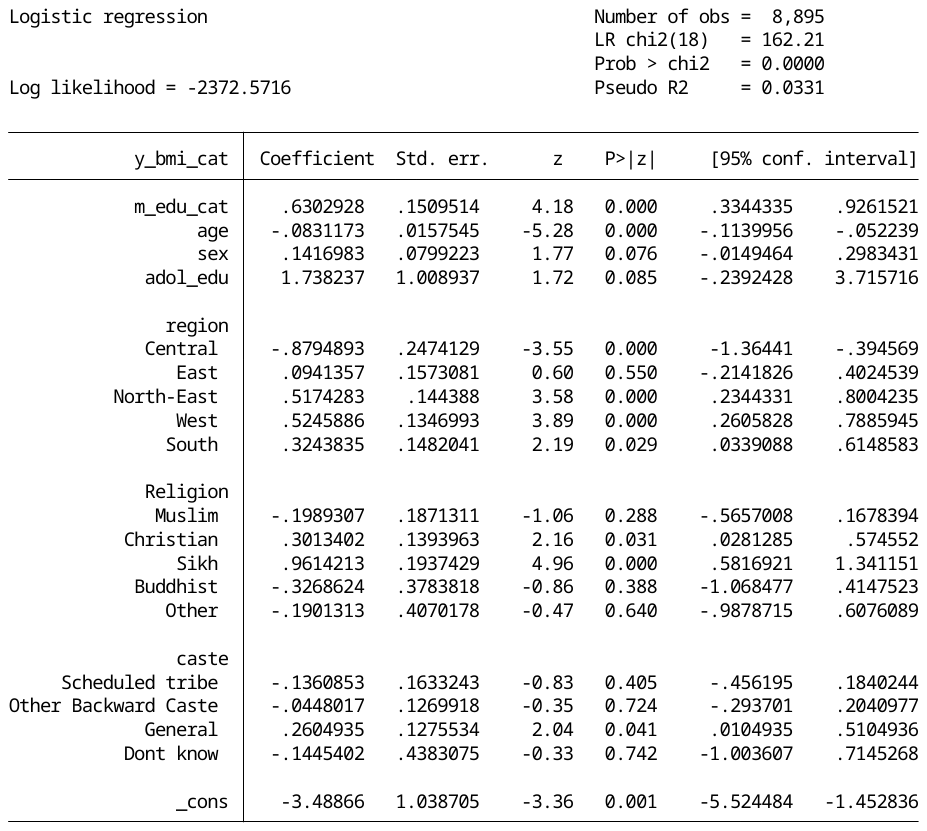

**Direct effect**

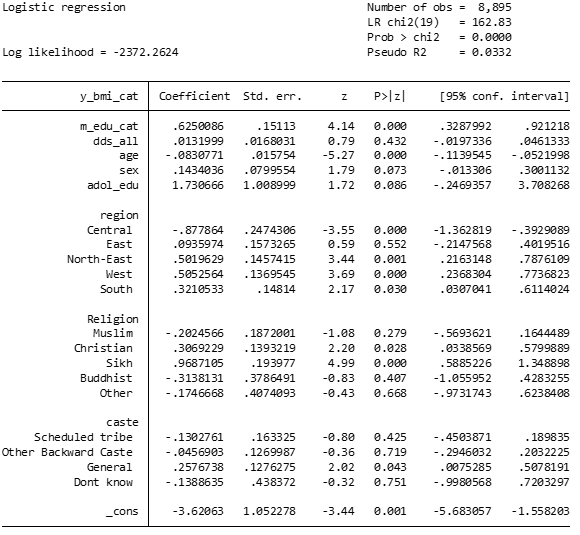

**Indirect effect**

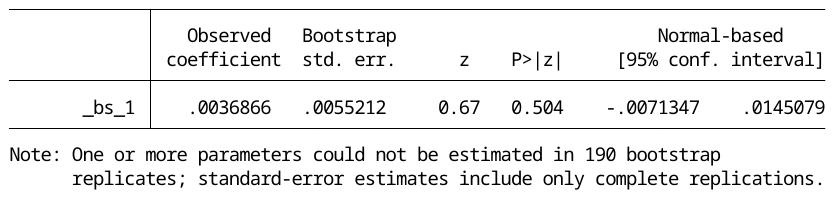

##### 3.1.2 Wealth index – DDS – overweight/obesity

**Exposure: w_index(binary); Mediator: dds_all (continuous); Outcomes: y_bmi_cat (binary)**

**Total effect**

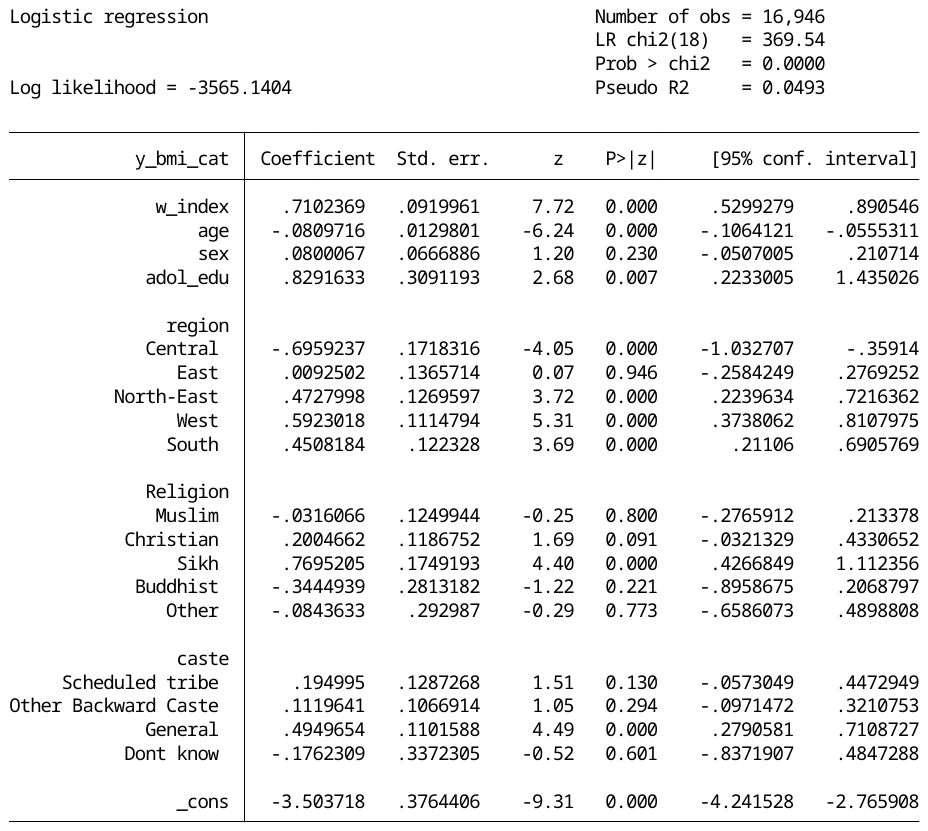

**Direct effect**

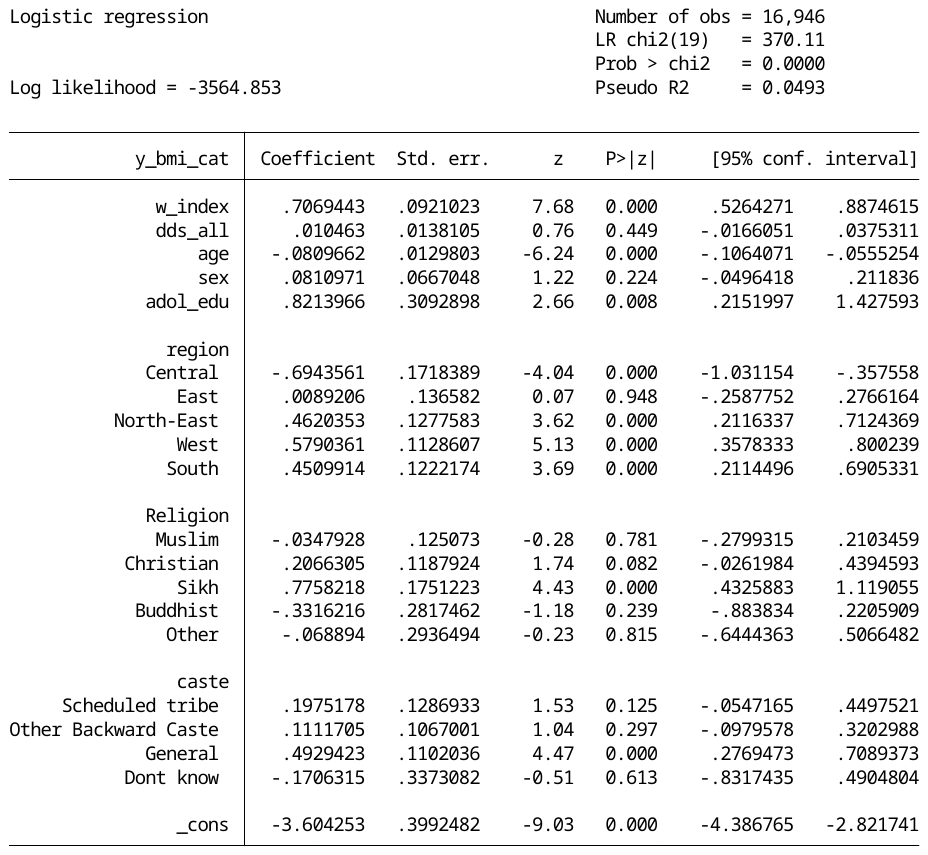

**Indirect effect**

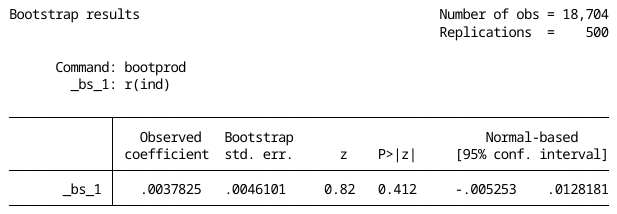

#### 4.2 Urban

##### 3.2.1 Maternal education – DDS – overweight/obesity

**Exposure: m_edu_cat (binary); Mediator: dds_all (continuous); Outcomes: y_bmi_cat (binary)**

**Total effect**

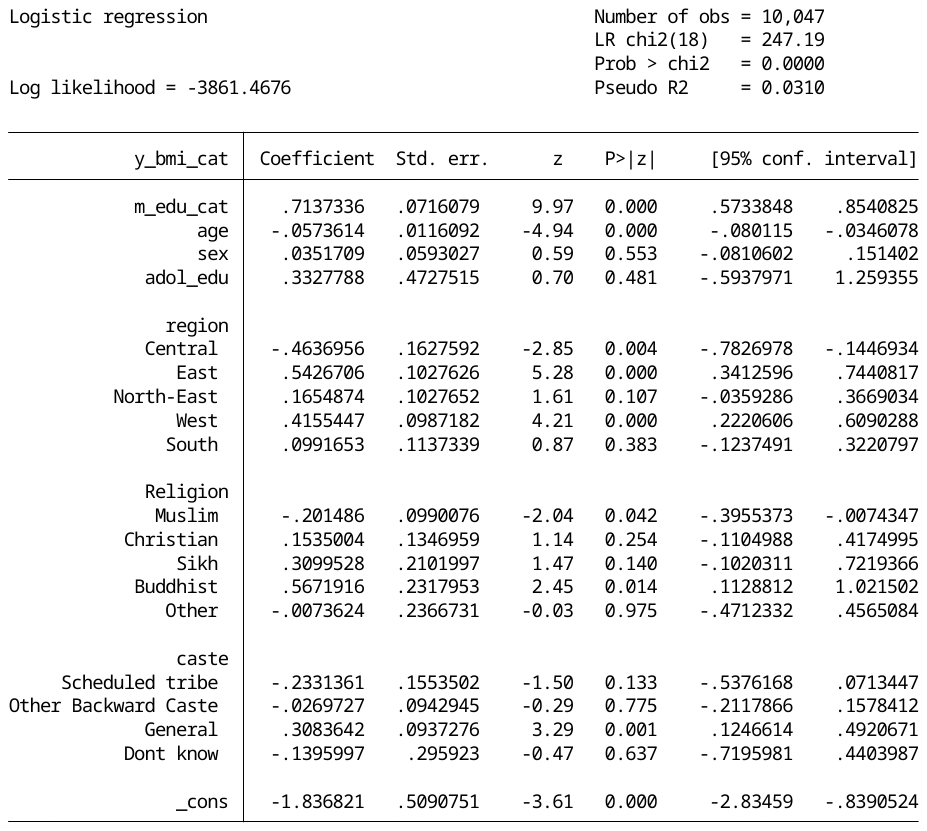

**Direct effect**

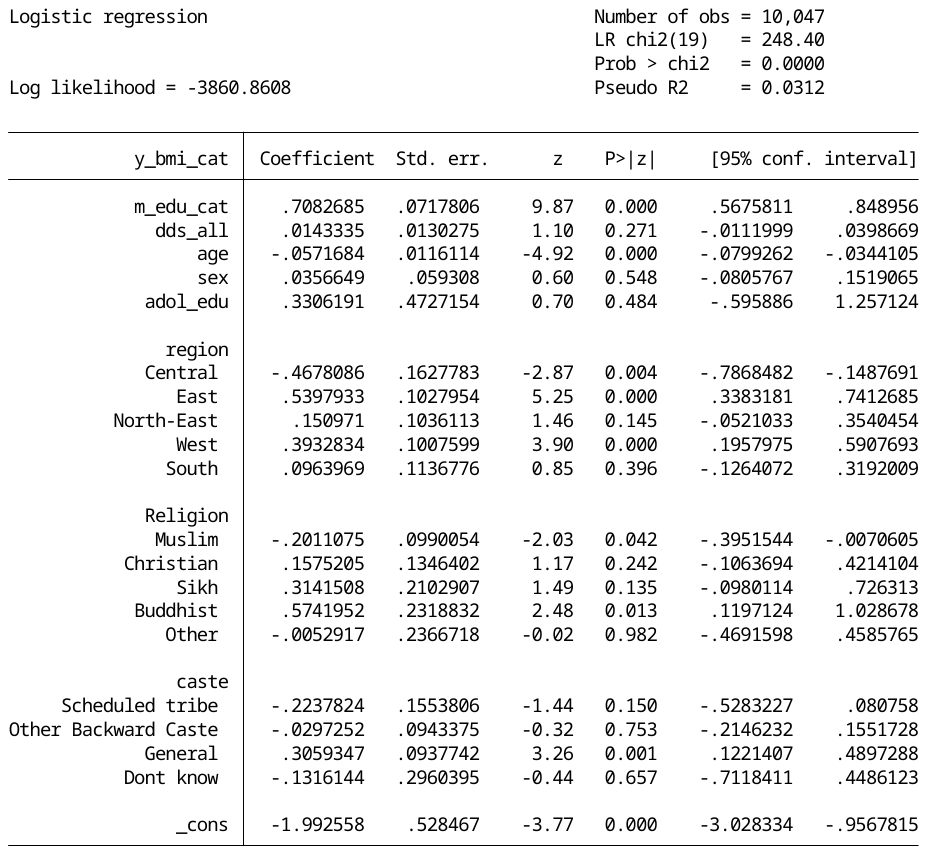

**Indirect effect**

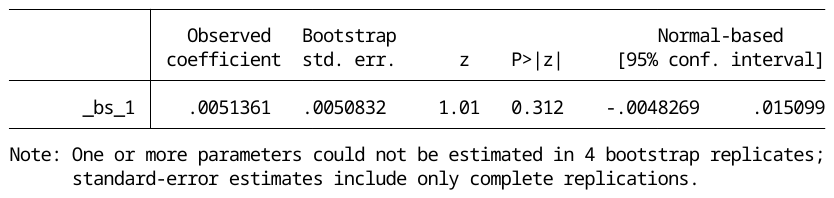

##### 3.2.2. Wealth index - DDS – overweight/obesity

**Exposure: w_index (binary); Mediator: dds_all (continuous); Outcomes: y_bmi_cat (binary)**

**Total effect**

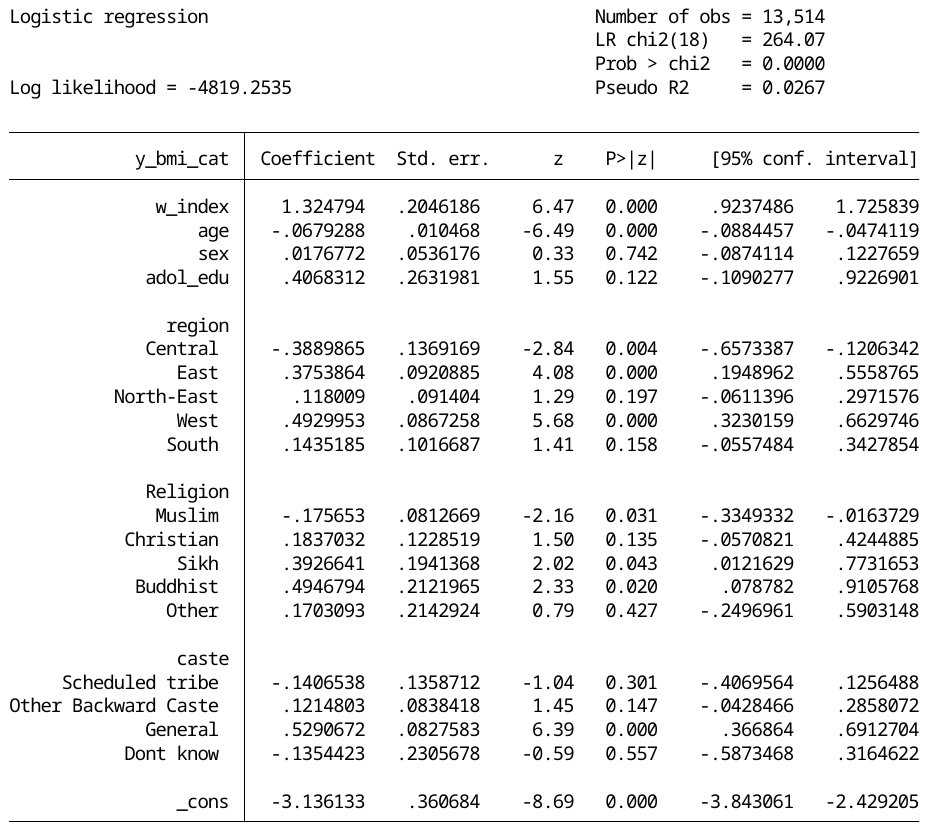

**Direct effect**

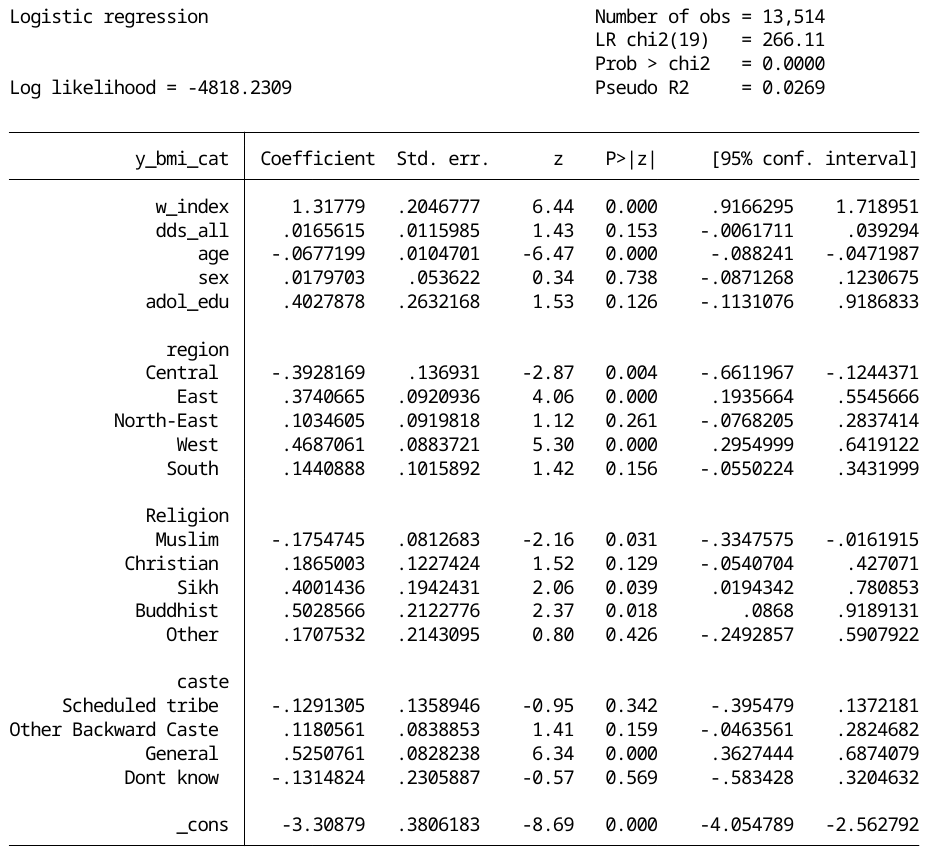

**Indirect effect**

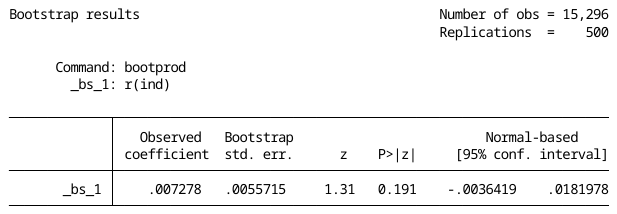

#### 4.3 Summary results of the base case

Table 1: Results from cross section mediation -RURAL

|  | **Coefficients** | **95% CI** | | **p-value** | **Proportion mediated** |
| --- | --- | --- | --- | --- | --- |
|  |  | **Low** | **High** |  |  |
| **Maternal education → Diet → Overweight/obesity** | | | | | |
| *Direct effects* | 0.625 | 0.329 | 0.921 | 0.000* | 0.63% |
| *Indirect effects* | 0.004 | -0.007 | 0.015 | 0.504 |  |
| *Total effect* | 0.630 | 0.334 | 0.926 | 0.000* |  |
| **Wealth → Diet → Overweight/obesity** | | | | | |
| *Direct effects* | 0.707 | 0.526 | 0.887 | 0.000* | 0.56% |
| *Indirect effects* | 0.004 | -0.005 | 0.013 | 0.412 |  |
| *Total effect* | 0.710 | 0.529 | 0.891 | 0.000* |  |

**Statistically significant as p<0.05*

Table 2: Results from cross section mediation -URBAN

|  | **Standard mediation** | | | | **Proportion mediated** |
| --- | --- | --- | --- | --- | --- |
|  | **Coefficients** | **95% CI** | | **p-value** |  |
|  |  | **Low** | **High** |  |  |
| **Maternal education → Diet → Overweight/obesity** | | | | | |
| *Direct effects* | 0.708 | 0.568 | 0.849 | 0.000* | 0.70% |
| *Indirect effects* | 0.005 | -0.005 | 0.015 | 0.312 |  |
| *Total effect* | 0.714 | 0.573 | 0.854 | 0.000* |  |
| **Wealth → Diet → Overweight/obesity** | | | | | |
| *Direct effects* | 1.318 | 0.917 | 1.719 | 0.000* | 0.53% |
| *Indirect effects* | 0.007 | -0.004 | 0.018 | 0.191 |  |
| *Total effect* | 1.325 | 0.924 | 1.726 | 0.000* |  |

**Statistically significant as p<0.05*

### 5. Robustness analyses

Table 3: Results from the robustness analyses- Rural population

| ***Exposure -> Mediator -> Outcome*** | | **Cross section mediation- Rural** | | | | |
| --- | --- | --- | --- | --- | --- | --- |
|  |  | **Coefficients** | **95% Confidence Intervals** | | **p-value** | **Proportion mediated** |
|  |  |  | **Lower** | **Higher** |  |  |
| ***Maternal education (binary) -> FVS (13 food groups) ->Obesity*** | | | | | | |
| Direct effects |  | 0.613 | 0.315 | 0.911 | 0.000* |  |
| Indirect effects |  | 0.016 | -0.016 | 0.048 | 0.322 | 2.54% |
| Total effect |  | 0.630 | 0.334 | 0.926 | 0.000* |  |
| ***Maternal education (binary) -> FVS_u (4 food groups) ->Obesity*** | | | | | | |
| Direct effects |  | 0.618 | 0.322 | 0.915 | 0.000* |  |
| Indirect effects |  | 0.013 | -0.004 | 0.029 | 0.129 | 2.06% |
| Total effect |  | 0.630 | 0.334 | 0.926 | 0.000* |  |
| ***Maternal education (binary) -> DDS (13 food groups) ->Obesity*** | | | | | | |
| Direct effects |  | 0.618 | 0.322 | 0.915 | 0.000* |  |
| Indirect effects |  | 0.010 | -0.005 | 0.024 | 0.213 | 1.59% |
| Total effect |  | 0.630 | 0.334 | 0.926 | 0.000* |  |
| ***Maternal education (binary) -> DDS_u (4 food groups) ->Obesity*** | | | | | | |
| Direct effects |  | 0.621 | 0.325 | 0.917 | 0.000* |  |
| Indirect effects |  | 0.009 | -0.005 | 0.023 | 0.198 | 1.43% |
| Total effect |  | 0.630 | 0.334 | 0.926 | 0.000* |  |
| ***Maternal education (continuous) -> DDS_all (17 food groups) ->Obesity*** | | | | | | |
| Direct effects |  | 0.097 | 0.072 | 0.121 | 0.000* |  |
| Indirect effects |  | 0.000 | -0.002 | 0.002 | 0.801 | 0.23% |
| Total effect |  | 0.097 | 0.072 | 0.121 | 0.000* |  |
| ***Wealth index (binary) -> FVS (13 food groups) ->Obesity*** | | | | | | |
| Direct effects |  | 0.693 | 0.512 | 0.875 | 0.000* |  |
| Indirect effects |  | 0.018 | -0.003 | 0.038 | 0.089 | 2.54% |
| Total effect |  | 0.710 | 0.530 | 0.891 | 0.000* |  |
| ***Wealth index (binary) -> FVS_u (4 food groups) ->Obesity*** | | | | | | |
| Direct effects |  | 0.690 | 0.509 | 0.871 | 0.000* |  |
| Indirect effects |  | 0.019 | 0.003 | 0.034 | 0.019 | 2.68% |
| Total effect |  | 0.710 | 0.530 | 0.891 | 0.000* |  |
| ***Wealth index (binary) -> DDS (13 food groups) ->Obesity*** | | | | | | |
| Direct effects |  | 0.697 | 0.516 | 0.878 | 0.000* |  |
| Indirect effects |  | 0.014 | -0.002 | 0.031 | 0.089 | 1.97% |
| Total effect |  | 0.710 | 0.530 | 0.891 | 0.000* |  |
| ***Wealth index (binary) -> DDS_u (4 food groups) ->Obesity*** | | | | | | |
| Direct effects |  | 0.687 | 0.506 | 0.868 | 0.000* |  |
| Indirect effects |  | 0.022 | 0.005 | 0.038 | 0.011 | 3.10% |
| Total effect |  | 0.710 | 0.530 | 0.891 | 0.000* |  |
| ***Wealth index (continuous) -> DDS_all (17 food groups) ->Obesity*** | | | | | | |
| Direct effects |  | 0.403 | 0.338 | 0.467 | 0.000* |  |
| Indirect effects |  | 0.001 | -0.004 | 0.005 | 0.784 | 0.16% |
| Total effect |  | 0.403 | 0.339 | 0.467 | 0.000* |  |

**Statistically significant at p<0.05*

Table 4 Results from the robustness analyses- Urban population

| ***Exposure -> Mediator -> Outcome*** | | **Cross section mediation- Urban** | | | | |
| --- | --- | --- | --- | --- | --- | --- |
|  |  | **Coefficients** | **95% Confidence Intervals** | | **p-value** | **Proportion mediated** |
|  |  |  | **Lower** | **Higher** |  |  |
| ***Maternal education (binary) -> FVS (13 food groups) ->Obesity*** | | | | | | |
| Direct effects |  | 0.679 | 0.536 | 0.822 | 0.000* |  |
| Indirect effects |  | 0.035 | 0.001 | 0.061 | 0.008 | 4.90% |
| Total effect |  | 0.714 | 0.573 | 0.854 | 0.000* |  |
| ***Maternal education (binary) -> FVS_u (4 food groups) ->Obesity*** | | | | | | |
| Direct effects |  | 0.707 | 0.567 | 0.848 | 0.000* |  |
| Indirect effects |  | 0.008 | 0.001 | 0.014 | 0.017 | 1.12% |
| Total effect |  | 0.714 | 0.573 | 0.854 | 0.000* |  |
| ***Maternal education (binary) -> DDS (13 food groups) ->Obesity*** | | | | | | |
| Direct effects |  | 0.695 | 0.554 | 0.836 | 0.000* |  |
| Indirect effects |  | 0.019 | 0.005 | 0.033 | 0.006 | 2.66% |
| Total effect |  | 0.714 | 0.573 | 0.854 | 0.000* |  |
| ***Maternal education (binary) -> DDS_u (4 food groups) ->Obesity*** | | | | | | |
| Direct effects |  | 0.702 | 0.562 | 0.843 | 0.000* |  |
| Indirect effects |  | 0.014 | 0.004 | 0.023 | 0.004 | 1.96% |
| Total effect |  | 0.714 | 0.573 | 0.854 | 0.000* |  |
| ***Maternal education (continuous) -> DDS_all (17 food groups) ->Obesity*** | | | | | | |
| Direct effects |  | 0.089 | 0.073 | 0.105 | 0.000* |  |
| Indirect effects |  | 0.001 | -0.001 | 0.002 | 0.401 | 1.11% |
| Total effect |  | 0.090 | 0.073 | 0.106 | 0.000* |  |
| ***Wealth index (binary) -> FVS (13 food groups) ->Obesity*** | | | | | | |
| Direct effects |  | 1.267 | 0.865 | 1.668 | 0.000* |  |
| Indirect effects |  | 0.055 | 0.031 | 0.079 | 0.000* | 4.15% |
| Total effect |  | 1.325 | 0.924 | 1.726 | 0.000* |  |
| ***Wealth index (binary) -> FVS_u (4 food groups) ->Obesity*** | | | | | | |
| Direct effects |  | 1.293 | 0.892 | 1.695 | 0.000* |  |
| Indirect effects |  | 0.026 | 0.012 | 0.040 | 0.000* | 1.96% |
| Total effect |  | 1.325 | 0.924 | 1.726 | 0.000* |  |
| ***Wealth index (binary) -> DDS (13 food groups) ->Obesity*** | | | | | | |
| Direct effects |  | 1.290 | 0.889 | 1.691 | 0.000* |  |
| Indirect effects |  | 0.033 | 0.014 | 0.052 | 0.001* | 2.49% |
| Total effect |  | 1.325 | 0.924 | 1.726 | 0.000* |  |
| ***Wealth index (binary) -> DDS_u (4 food groups) ->Obesity*** | | | | | | |
| Direct effects |  | 1.283 | 0.881 | 1.684 | 0.000* |  |
| Indirect effects |  | 0.036 | 0.019 | 0.053 | 0.000* | 2.72% |
| Total effect |  | 1.325 | 0.924 | 1.726 | 0.000* |  |
| ***Wealth index (continuous) -> DDS_all (17 food groups) ->Obesity*** | | | | | | |
| Direct effects |  | 0.572 | 0.491 | 0.654 | 0.000* |  |
| Indirect effects |  | 0.001 | -0.003 | 0.006 | 0.552 | 0.24% |
| Total effect |  | 0.574 | 0.493 | 0.655 | 0.000* |  |

**Statistically significant at p<0.05*
